# Platelet Factor 4 Antibody Persistence and Long-term Pathogenicity in Vaccine-induced Immune Thrombotic Thrombocytopenia

**DOI:** 10.1101/2025.11.24.25340592

**Authors:** Adam Kanack, Emily Mauch, Guillaume Roberge, Noah Splinter, Krishna Gundabolu, Geoffrey D. Wool, Gemlyn George, Mouhamed Yazan Abou-Ismail, Kristi J. Smock, David L. Green, Jonathan Coker, Mindy C. Kohlhagen, David L. Murray, Anand Padmanabhan

**Affiliations:** Department of Laboratory Medicine and Pathology, Mayo Clinic, Rochester, MN; Centre d’Excellence des Maladies Vasculaires, Centre Hospitalier Universitaire de Québec, Hôpital Saint-François d’Assise, Université Laval, Québec, Canada; Fred & Pamela Buffett Cancer Center, University of Nebraska Medical Center College of Medicine, Omaha, NE; Department of Pathology, University of Chicago, Chicago, IL; Department of Medicine, University of Colorado, Aurora, CO; Department of Medicine, University of Utah, Salt Lake City, UT; Department of Pathology and ARUP Laboratories, University of Utah, Salt Lake City, UT; NYU Langone Health, New York, NY

## Abstract

Rarely, recipients of adenoviral vector-based vaccines experience a severe thrombotic thrombocytopenic condition referred to as vaccine-induced immune thrombotic thrombocytopenia (VITT). VITT is a transient prothrombotic process, although recent data suggests that VITT anti-platelet factor 4 (PF4) antibodies are more persistent than antibodies seen in heparin-induced thrombocytopenia. Whether anti-PF4 antibody persistence in VITT is related to the continued persistence of antibody clones from the acute phase or the development of novel antibodies is unclear. To study this, acute and follow-up samples were obtained from six Ad26.COV2.S-associated VITT patients, with a median time to follow-up of 244 days from acute presentation (Range, 114-664 days). Upon affinity-enrichment of antibodies, mono/oligoclonal PF4/heparin-reactive anti-PF4 antibodies were observed despite negative results in serum protein electrophoresis and the more sensitive “Mass-Fix” technique. This finding distinguishes VITT from monoclonal gammopathy of thrombotic significance where monoclonal antibodies are observed in native sera. Anti-PF4 antibody abundance decreased over time, with no evidence of novel anti-PF4 antibody production after acute presentation. Although previous studies indicate a stereotypical pairing of VITT antibodies with *lambda* light chains, one VITT patient produced anti-PF4 antibodies with a *kappa* light chain, suggesting immunological heterogeneity. While none of these six antibodies caused long-term thrombocytopenia or thrombosis, platelet-activating anti-PF4 antibodies were seen four years after the acute event in an additional ChAdOx1 nCoV-19-associated VITT patient. These antibodies continued to cause chronic low-grade thrombocytopenia, highlighting the potential for long-term sequelae in what is generally viewed as a transient thrombotic thrombocytopenic syndrome.

## Introduction

The emergence of severe acute respiratory syndrome coronavirus 2 (SARS-CoV-2) in 2019 led to a broad vaccination effort aimed at controlling the morbidity and mortality of COVID-19. In rare cases, recipients of ChAdOx1 nCoV-19 (AstraZeneca) and Ad26.COV2.S (Janssen Johnson & Johnson) vaccines have experienced a life-threatening adverse reaction termed vaccine-induced immune thrombotic thrombocytopenia (VITT), typically presenting 8-14 days post-vaccination and characterized by thrombocytopenia and thrombosis, with a high associated rate of morbidity and mortality.^1^ Underpinning the thrombotic and thrombocytopenic complications in VITT is the production of anti-platelet factor 4 (PF4) antibodies, which successively bind to PF4 and IgG Fc receptors (i.e., FcγRIIa) on the platelet surface, and stimulate platelet activation^2^. This anti-PF4 antibody-mediated mechanism of platelet activation in VITT is similar to that of the well-studied thrombotic disorder, heparin-induced thrombocytopenia (HIT), and the more recently described thrombotic disorder, monoclonal gammopathy of thrombotic significance (MGTS). MGTS is defined by the presence of a monoclonal antibody (M-protein) diagnostically confirmed to be a platelet-activating anti-PF4 antibody^3,4^.

Here, we endeavored to better define the immunological responses governing the production of anti-PF4 antibodies in the context of VITT and to further differentiate these immunological responses from the more attenuated anti-PF4 antibody production that is a hallmark of HIT.^3,5^ In this study, affinity-purified anti-PF4 antibodies, as well as immunoglobulin G (IgG) from the entire serum antibody repertoire, were evaluated in the acute and follow-up samples of six Ad26.COV2.S VITT patients and samples from one patient with ChAdOx1 nCov-19-associated VITT. Results demonstrate that persistence of the acute anti-PF4 antibody, rather than the generation of novel anti-PF4 antibodies, underlies antibody persistence in VITT. Despite the presence of clonally-restricted anti-PF4 antibodies in VITT, mono/oligoclonal antibodies are not detected in conventional testing for M-proteins using techniques such as serum protein electrophoresis (SPEP) and “Mass-Fix”.^6^ We further demonstrate that in the ChAdOx1 nCov-19-associated VITT patient, a persistent anti-PF4 antibody continues to cause thrombocytopenia more than 4 years after the acute event.

## Methods

To evaluate each patient’s total antibody repertoire at acute presentation and their clonal anti-PF4 antibody profiles at the acute and convalescent stages, multiple patient samples were obtained from six individuals with Ad26.COV2.S-associated VITT. Acute samples were defined as samples drawn within 40 days of symptom onset. Numerous follow-up samples from the ChAdOx1 nCov-19-associated VITT patient were studied, as acute samples were not available. Anti-PF4 antibodies were immuno-enriched from all patients using PF4-treated heparin sepharose beads, and additionally with covalently linked PF4 beads in the ChAdOx1 nCov-19-associated VITT patient. Isolated anti-PF4 antibodies and serum samples were evaluated in PF4-polyanion HIT ELISAs and functional, platelet-based assays as previously described^3^. Sera and isolated anti-PF4 antibodies were subjected to liquid chromatography-electrospray ionization- quadrupole time-of-flight mass spectrometry (LC-ESI-QTOF MS) as previously described^3^. Serum protein electrophoresis (SPEP) and matrix-assisted laser desorption ionization time-of-flight mass spectrometry (“Mass-Fix”) were performed to assess the presence of M-proteins in native serum. Detailed methods are provided in the *Supplementary Appendix*. The Institutional Review Board of Mayo Clinic approved the research studies.

## Results

The Ad26.COV2.S VITT cohort included four male and two female patients (**Table S1**). Median time from Ad26.COV2.S vaccination to onset of symptoms was 10.5 days (Range, 8 – 14 days). All patients presented with thrombocytopenia, with a median platelet count of 25,500/µL (Range, 9000 – 126,000/µL), and thrombosis. The most common thrombotic location was splanchnic vein thrombosis in four patients and cerebral venous sinus thrombosis (CVST) in three patients. Two patients developed both splanchnic vein thrombosis and CVST. During acute hospitalization, all patients received high-dose intravenous immunoglobulin G (IVIg) and a direct thrombin inhibitor, while five of the six patients also received steroid treatment. All patients’ follow-up treatments included anticoagulation with direct oral anticoagulants (DOACs). Two patients received additional treatment with IVIg for recurrent thrombocytopenia, and one patient with persistent thrombocytopenia was treated with IVIg, rituximab, and therapeutic plasma exchange. None of the patients experienced breakthrough thrombosis after anticoagulation treatment was initiated (**Table S1**).

The median age of the latest follow-up sample was 244 days from acute presentation (Range, 114-664 days). High-resolution mass spectrometry profiling (liquid chromatography-electrospray ionization quadrupole time-of-flight mass spectrometry, LC-ESI-QTOF-MS) of acute patient sera and anti-PF4 immuno-enriched samples demonstrated monoclonal anti-PF4 antibodies in patients 2, 4, 5, and 6 (**Figs 1B, D-F**). Patients 1 and 3 showed biclonal and triclonal anti-PF4 antibody responses (**Figs 1A, C**). Monoclonal proteins (M-proteins) matching the mass of the respective immuno-enriched anti-PF4 antibodies were identifiable above the polyclonal background in the sera of patients 2, 5, and to a lesser degree, patient 6 (**Figs 1B, E, and F**). A clear M-protein noted in the spectra of patient 6 corresponded to rituximab therapy. As expected, a polyclonal antibody profile was observed from the sera of a healthy donor control, and anti-PF4 antibodies were not immuno-enriched from this sample (**Fig S1**).

**Figure 1.**
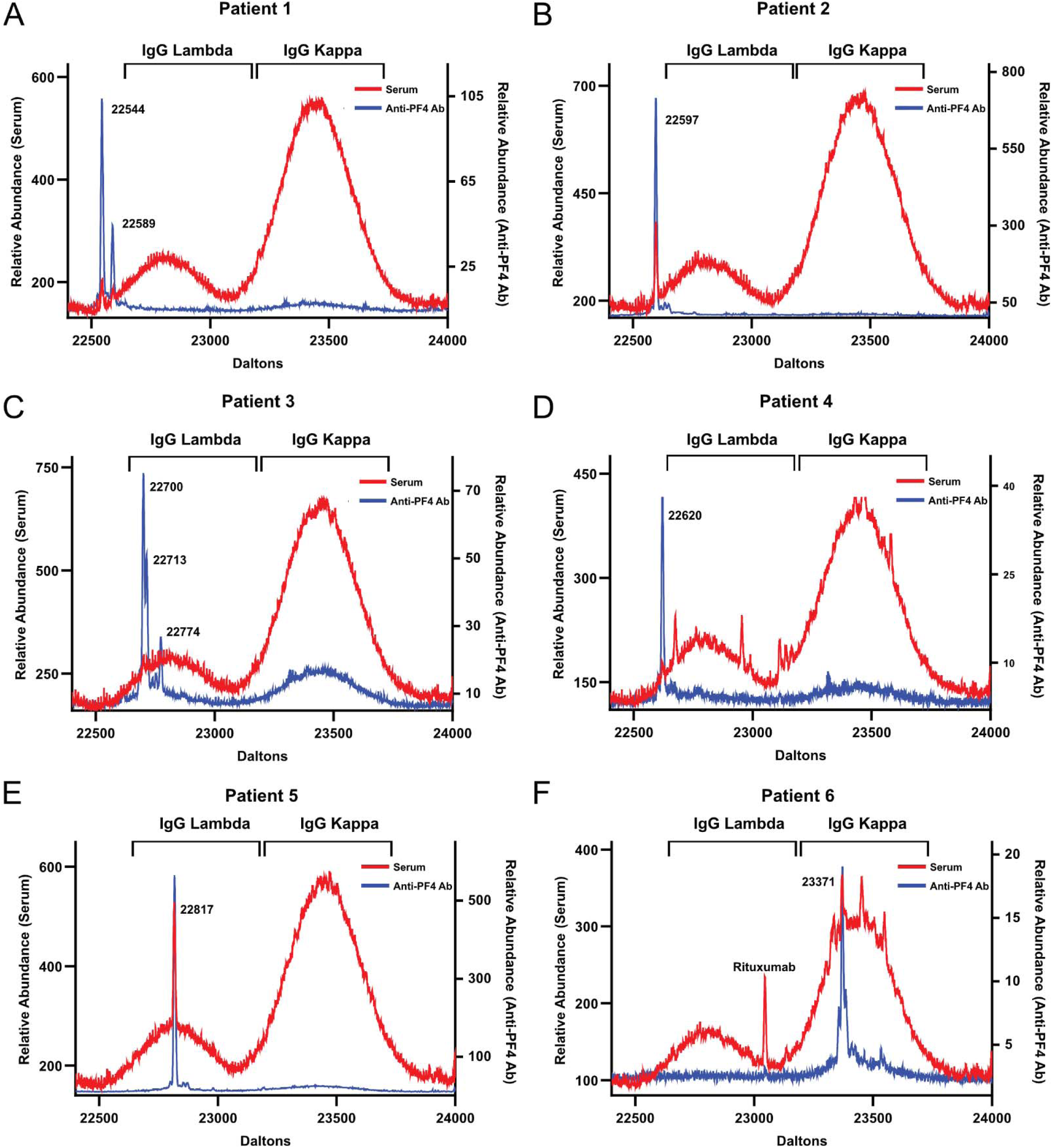
VITT patients displayed a polyclonal serum antibody profile and clonally restricted anti-PF4 antibody spectra. **(A-F)** LC-ESI-QTOF MS calculated mass spectra (Daltons), based on [M + 12H]^12+^ light chain charge state associated with IgG heavy chains of acute sera (red), or anti-PF4 antibodies (blue) from acute sera from patients 1-6 is shown. Prominent immunoglobulin peak values are displayed on each graph. In Patient 6 (**F**), an additional peak corresponds to rituximab, which the patient received before the acute sample blood draw. A mass range of 22400 – 24000 Daltons is shown for each panel.

At the time of the last follow-up sample (Median, 184 days; Range, 90 – 646 days), IgG antibodies positive in PF4/polyanion HIT ELISA testing were detected in the sera of 5/6 VITT patients (**Table S2**). After immuno-enrichment of each patient’s last follow-up sample, affinity-purified anti-PF4 antibodies were detected by HIT ELISA in all six samples (**Fig 2A-F**). Four of six affinity-purified anti-PF4 antibody samples stimulated platelet activation at least at one follow-up time point (patients 1, 2, 3, and 6; **Fig 2A, B, C & F**). Remarkably, Patient 6 produced an anti-PF4 antibody with an IgG *kappa* light chain (**Fig 1F, 2F, and Fig S2**). In contrast, the remaining five VITT patients produced anti-PF4 antibodies with IgG *lambda* light chains, aligning with previous VITT studies examining antibody clonality (**Figs 1A-E and 2A-E**).^7,8^ In all patients, the relative abundance of the immuno-enriched anti-PF4 antibodies was lower in follow-up samples relative to acute samples (**Fig 2A-F**). High-resolution mass spectrometry of anti-PF4 antibodies did not reveal any novel anti-PF4 clones in any of the six VITT patients (**Fig 2A-F**). Prior research suggests the presence of stereotyped clonotypic anti-PF4 antibodies in VITT exclusively with *lambda* light chains^8^. However, one of our VITT patients’ monoclonal anti-PF4 antibodies contained *kappa* light chains, highlighting the complexity and immune heterogeneity of this disorder.

**Figure 2.**
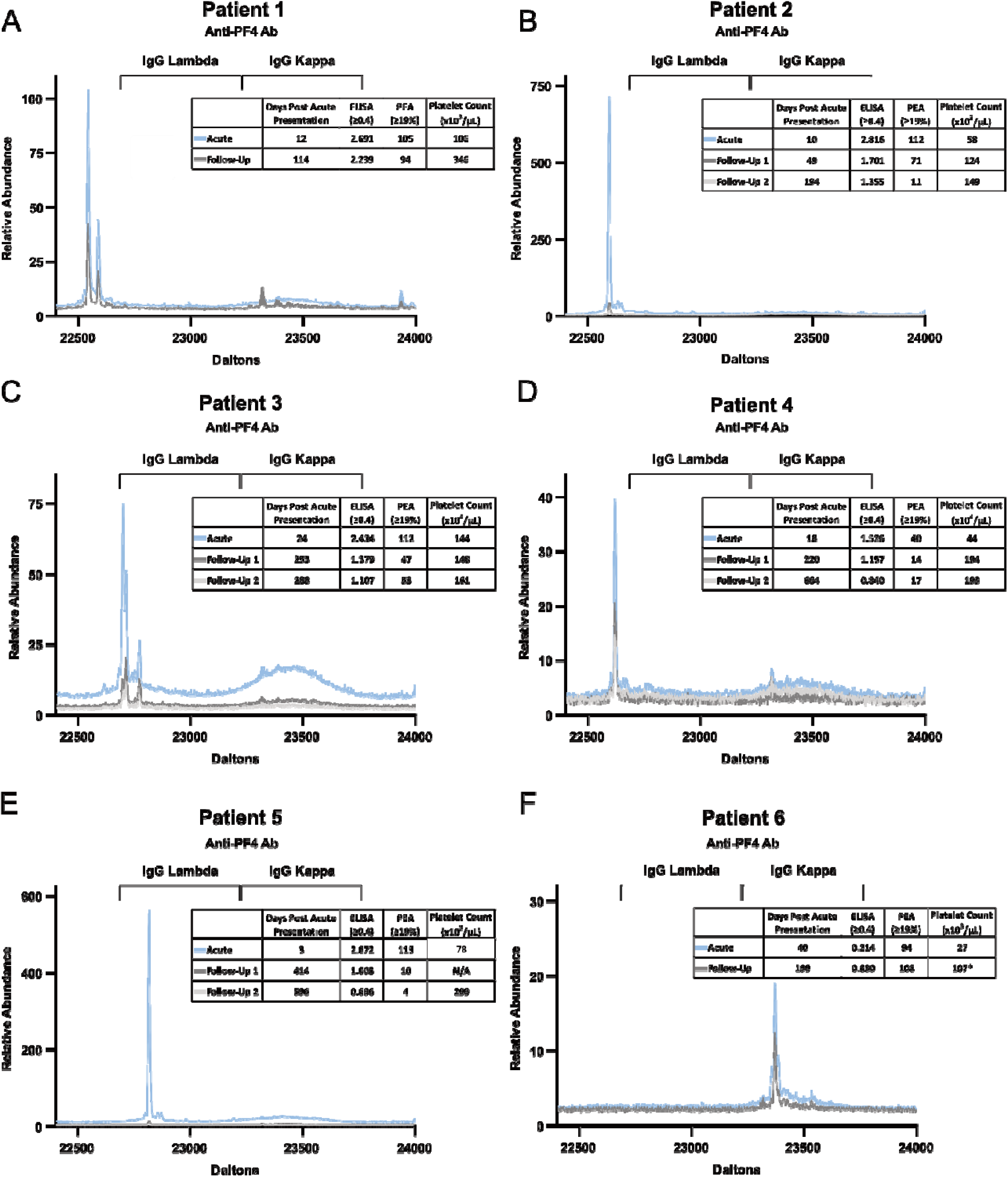
VITT antibodies are highly persistent. (**A-F**). LC-ESI-QTOF MS calculated mass spectra (Daltons) based on [M + 12H]^12+^ light chain charge state associated with IgG heavy chain of acute (blue) and convalescent (dark gray, light gray) samples in patients 1-6, are shown. PF4/polyanion ELISA, PF4-dependent P-Selectin Expression (PEA), platelet counts, and time after acute presentation are displayed in the inset table of each subfigure. A mass range of 22400 – 24000 Daltons is shown for each panel. The asterisk indicates that no platelet count was available on the date of follow-up of this sample; the platelet count presented was obtained within a week of the tested sample.

SPEP and a recently-deployed mass spectrometry-based diagnostic test for M-proteins, matrix-assisted laser desorption ionization time-of-flight mass spectrometry (“Mass-Fix”)^6^ were performed to evaluate whether these diagnostic tools would detect mono/oligoclonal anti-PF4 VITT antibodies in sera. Sufficient volume availability was limited to patients 3, 5, and 6. No diagnosable M-spikes were observed in SPEP testing (**Figs 3A-C**) or Mass-Fix (**Figs 3D-F**), even though these patients produced monoclonal (patients 5 and 6) or oligoclonal (patient 3) anti-PF4 antibodies.

**Figure 3.**
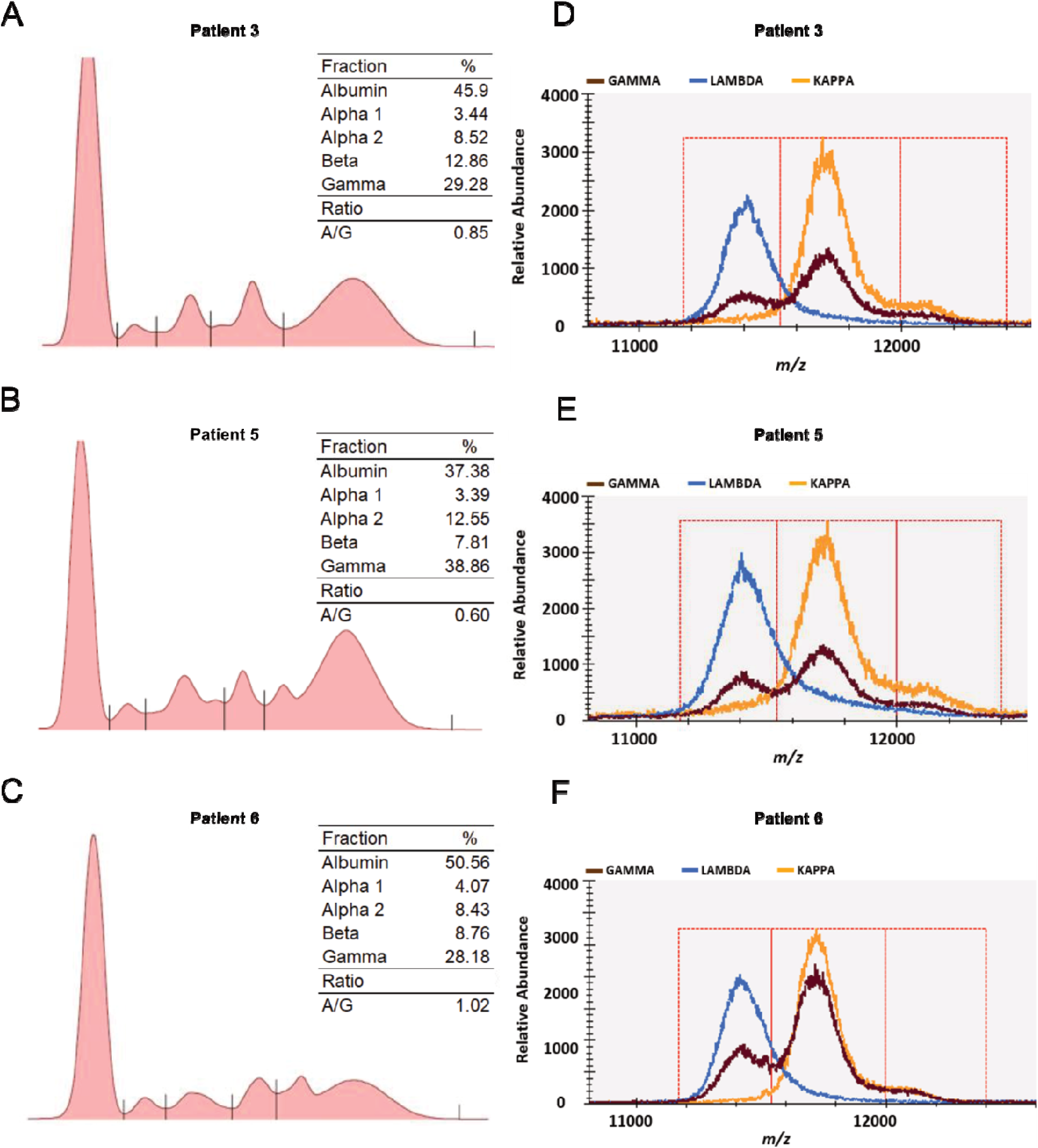
VITT patients show no evidence of monoclonal gammopathies by serum protein electrophoresis (SPEP) or Mass-Fix. **(A-C)** Relative densitometry quantification results for the major plasma components are presented as a percentage of the total protein signal. The ratio of albumin to gamma-globulin is also displayed. No mono/oligoclonal gammopathies were identified. (**D-F**) The +2 charge state spectra of the acute serum samples for Patients 3, 5 and 6 for IgG heavy chain (GAMMA) in brown, lambda containing immunoglobulins (Igs) (LAMBDA) in blue, and kappa containing Igs (KAPPA) in orange are displayed in the Mass-Fix test. The x-axis depicts mass-charge (m/z), and the y-axis presents the relative abundance of identified antibodies.

The ChAdOx1 nCov-19-associated VITT patient was a male in his 50s with a history of hypertension, and hypercholesterolemia who twenty days post ChAdOx1 nCoV19 vaccination developed right lower limb proximal deep vein thrombosis, segmental pulmonary embolisms, cerebral vein thrombosis of the left sigmoid sinus extending to the jugular vein, and infrarenal aortic and left popliteal artery thrombi (described previously^9,10^, and **Figure 4A**). Upon admission, a decreased platelet count of 58 x 10^3^/µL and an elevated D-dimer level of 16,561 ng/mL FEU were observed. A diagnosis of VITT was subsequently made following positivity in PF4-polyanion HIT ELISA and PF4-enhanced Serotonin Release Assay (PF4-SRA) testing. Initial treatment included argatroban (2.5 µg/kg/min), intravenous immunoglobulin G (IVIg, 1 g/kg x 2 doses), and prednisone (1 mg/kg). The patient required additional IVIg infusions and popliteal artery recanalization during his hospitalization and was transitioned to long-term rivaroxaban (20 mg daily) prior to discharge (**Figure 4A**).

**Figure 4.**
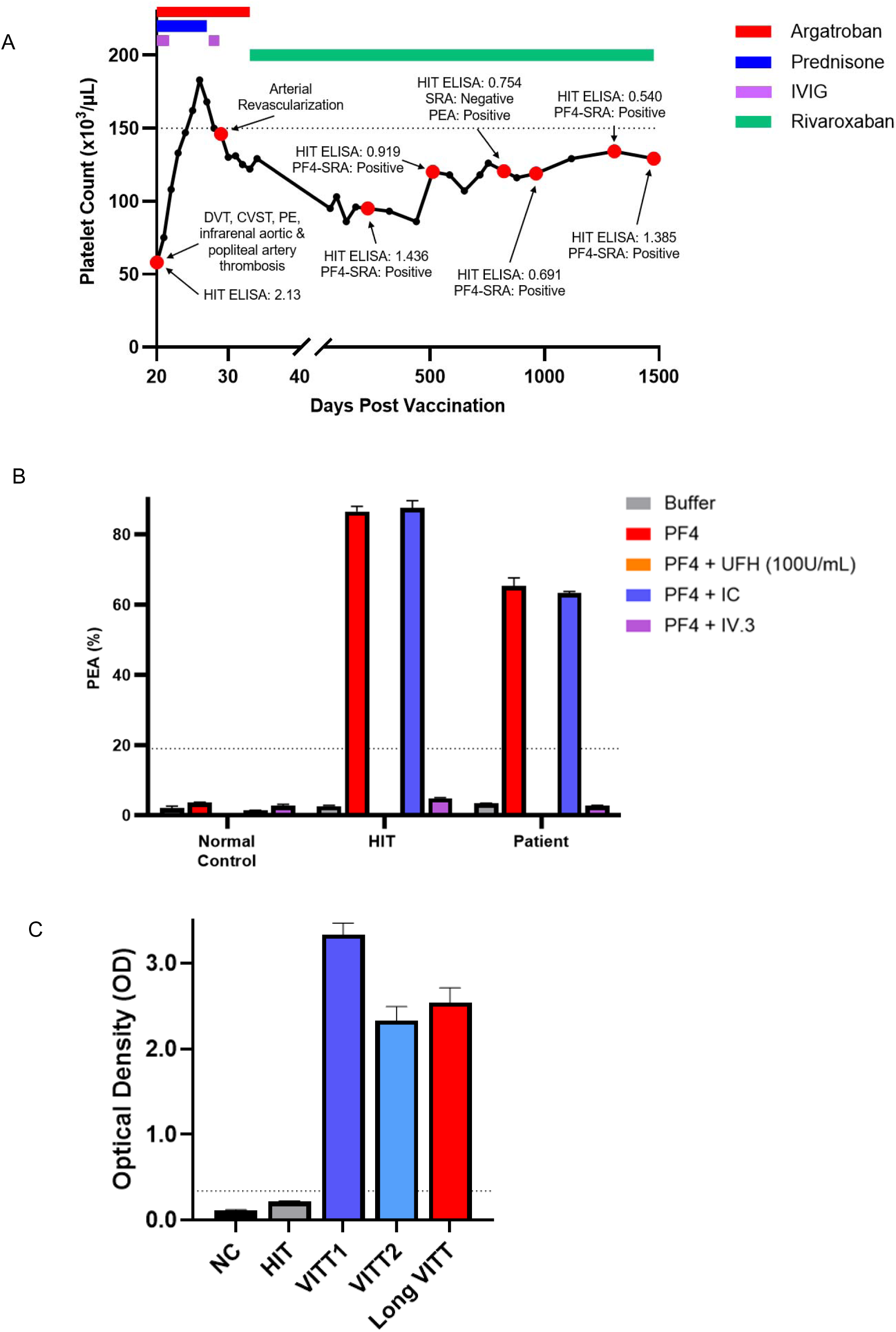
Patient antibodies are highly persistent, ELISA positive, and mediate PF4-dependent platelet activation. **(A)** Patient platelet counts (black circles) are displayed starting from the time of vaccination. Red circles denote major clinical events or diagnostic testing results. Patient treatment and medications are denoted at the top of the graph. IVIg represents therapy with intravenous immunoglobulin G. **(B)** Normal serum (Normal Control), HIT patient serum (HIT), or patient serum were evaluated in the PF4-dependent P-selectin expression assay (PEA). Testing conditions included buffer-treated platelets, PF4-treated platelets, PF4-treated platelets with unfractionated heparin (UFH, 100 U/mL), PF4-treated platelets with murine antibody isotype control (IC, 2 µg/mL), or PF4-treated platelets with Fc gamma region receptor II-a (FcγRIIa) blocking murine monoclonal antibody IV.3 (2 µg/mL). **(C)** Patient and two Ad26.COV2.S-associated VITT sera recognize uncomplexed PF4 targets. Dotted lines represent the positive cut-offs/lower limit of the reference normal range of the respective tests.

In follow-up that has lasted more than four years after the acute VITT hospitalization, he has remained chronically thrombocytopenic (platelet count range: 86 to 134 x 10^3^/µL) with persistently positive PF4-polyanion ELISA and PF4-dependent functional tests (**Figure 4A).** He has not experienced recurrent thrombotic events or required subsequent IVIg treatment while continuing rivaroxaban therapy. The patient’s serum was assessed for platelet-activating anti-PF4 antibodies using heparin- and PF4-based functional platelet assays to further characterize the pathogenic antibody. In congruence with classical VITT serology, patient serum activated platelets in the presence of PF4 (PEA & PF4-SRA; **Figure 4A-B**) but not heparin (conventional SRA; **Figure 4A**). Additional functional testing in the presence of a high concentration of heparin or the Fc receptor-blocking monoclonal antibody IV.3 demonstrated a reactivity pattern identical to that of a platelet-activating anti-PF4 antibody (**Figure 4B**). The patient also tested positive in a VITT-specific ELISA that uses uncomplexed PF4 as a target (**Figure 4C**).

To provide additional insight into the persistence of the patient’s anti-PF4 antibodies, a mass-spectrometry-based assessment of the patient’s native serum and anti-PF4 antibody repertoires was performed. Unlike in MGTS, serum protein electrophoresis did not reveal a monoclonal gammopathy (**Figure 5A**). Similarly, Mass-Fix testing of patient serum, was reported to be negative for a monoclonal antibody (**Supplementary Figure S3**). While LC-ESI-QTOF mass spectrometry of native serum revealed a polyclonal antibody profile, two monoclonal spikes were seen above the polyclonal background (**Figure 5B**: 22,668Da, 23,491Da). The patient’s serum was immuno-enriched for anti-PF4 antibodies using PF4-coupled beads (as described in the *Supplementary Appendix*). The isolated eluate demonstrated binding to PF4-polyanion targets by HIT ELISA (**Figure 5C**) and stimulated the activation of platelets in PF4-based functional platelet assays (**Figure 5D**) confirming the isolation of functional anti-PF4 antibodies. High resolution LC-ESI-QTOF mass spectrometry of the isolated anti-PF4 antibodies displayed a monoclonal IgG *lambda* light chain with near-identical mass to one of the two prominent monoclonal proteins observed in the sera (**Figure 5E vs. 5B:** 22,666Da vs 22,668Da). To confirm the presence of an anti-PF4 antibody in the patient’s serum using an orthogonal strategy, an additional immuno-enrichment of anti-PF4 antibodies from native serum was performed using PF4-treated heparin sepharose beads. LC-ESI-QTOF MS of this anti-PF4-polyanion-based enrichment again demonstrated the presence of a monoclonal IgG λ light chain with a nearly identical mass to the anti-PF4 antibody previously isolated using PF4-coupled beads and matched the monoclonal protein profile observed with the patient’s serum (**Supplementary Figure S4**, 22,665Da in PF4-heparin bead eluate vs. 22,666 Da in PF4-bead eluate (**2E**) vs. 22,668Da in serum, **5B**).

**Figure 5.**
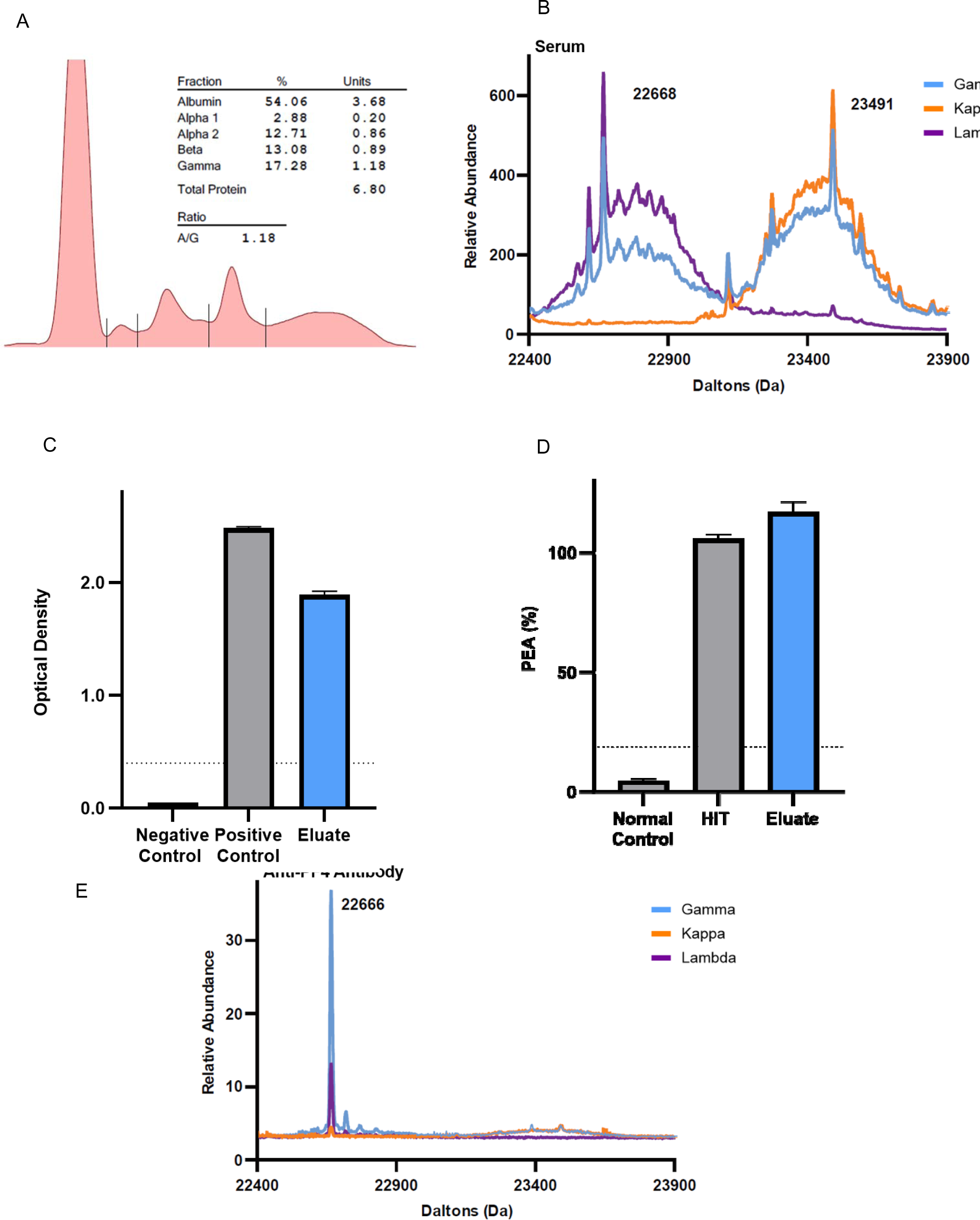
Mass spectrometry profiles of native patient serum and anti-PF4 antibodies. **(A)** Serum protein electrophoresis of the patient’s serum displays total protein quantification in percentage (%) and units (g/dL). The albumin-to-globulin ratio is presented as A/G. No monoclonal bands were seen. **(B)** LC-ESI-QTOF MS +11 deconvoluted charge state spectra (Daltons) of patient serum light chains are shown. **(C)** Isolated anti-PF4 antibodies eluted from PF4-coupled beads bind PF4-polyanion complexes (PF4 IgG, Immucor) and **(D)** activate PF4-treated platelets. The dotted line represents the positive cut-offs of the respective assays. **(E)** LC-ESI-QTOF MS results of the isolated anti-PF4 antibodies in the +11 deconvoluted charge state spectra (Daltons) are shown. **(B)** and **(E)** Display light chain distributions associated with an IgG heavy chain (*Gamma*) in blue, kappa-containing Igs (*Kappa*) in orange, and lambda containing Igs (*Lambda*) in purple. Values indicated on the graph represent the mass (Da) of the most prominent immunoglobulin light chains.

## Discussion

Previous long-term follow-up studies of VITT patients have demonstrated a disconnect in anti-PF4 antibody persistence, whereby solid-phase HIT ELISA positivity persists significantly longer than positivity in platelet-based functional testing.^2,11,12^ One hypothesis to explain this discordance is epitope spreading, whereby, over time, the specificity of anti-PF4 antibodies may decrease for PF4 epitopes elicited by its interactions with the platelet surface while becoming enhanced for the PF4 epitopes elicited by the polyanions utilized in diagnostic HIT ELISAs. Alternatively, the plasma concentration of anti-PF4 antibodies may gradually decrease, resulting in insufficient anti-PF4 antibodies to stimulate platelet FcγRIIa cross-linking and platelet activation.

Based on the study of our cohort, clonally restricted anti-PF4 antibodies associated with VITT can persist for years with no evidence of novel anti-PF4 antibody production after acute presentation. The decrease in the relative abundance of anti-PF4 antibodies noted in this study, rather than a shift in antibody specificity (which could result in epitope spreading and the production of non-pathogenic anti-PF4 antibodies), likely underlies the transient prothrombotic state in VITT. Unlike in MGTS, where monoclonal anti-PF4 antibodies are produced in the context of an easily recognized monoclonal gammopathy, these VITT cases highlight the production of monoclonal/oligoclonal anti-PF4 antibodies in the absence of a detectable M-protein in serum-based testing (i.e., the mono/oligoclonal antibody cannot be detected without evaluation of affinity-enriched anti-PF4 antibodies). Whether these persistent anti-PF4 clones in convalescent patients can re-activate and cause thrombosis in settings such as infection or inflammation requires further follow-up and observation. Since the conclusion of this study, additional follow-up information was available on five of the six Ad26.COV2.S-associated VITT patients: All five were doing well without recurrence of thrombosis, and all but one, who was on apixaban, had stopped anticoagulation therapy. PF4/polyanion ELISA testing results were available in four patients with a median time of 1251 days (>3 years) after acute presentation (range 1170-1466 days). Three of the four patients continued to test positive for anti-PF4 antibodies (range 0.406-1.427 OD) further confirming the highly persistent nature of VITT antibodies.

As highlighted in the ChAdOx1 nCoV-19-associated VITT case, these antibodies can cause long-term (> 4 years) thrombocytopenia and persistent positivity in PF4 antigen-based and functional tests. Mirroring the Ad26.COV2.S-associated VITT patients, conventional diagnostic tests for monoclonal gammopathy, such as the SPEP and the recently deployed more sensitive Mass-Fix test, did not detect a monoclonal anti-PF4 antibody. The patient’s antibody/ presentation displayed aspects of VITT and MGTS. Like most VITT antibodies, this patient’s monoclonal antibody was an immunoglobulin G (IgG) antibody with lambda light chains^7,8^. Unlike most VITT antibodies, this patient’s monoclonal anti-PF4 antibody caused thrombocytopenia for several years, similar to MGTS antibodies^3,4,13^. However, unlike patients with MGTS, this VITT patient tested negative in conventional testing for monoclonal gammopathy of undetermined significance, suggesting the concentration of pathogenic antibodies was below the detection threshold of currently deployed diagnostic methodologies. Importantly, the use of anticoagulation was effective in preventing additional thrombosis. Whether additional treatments are necessary to address the persistent pathogenic anti-PF4 antibody, such as those used recently in MGTS^13,14^ requires further evaluation.

## Data Availability

All data produced in the present study are available upon reasonable request to the authors

## Acknowledgments

This work was supported, in part, by the National Institutes of Health grants HL171911 (AJK) and HL158932 (AP).

## Authorship

AJK, EEM, and AP collaborated on designing research studies. AJK, EEM, GR and AP collaborated on the first draft of the manuscript, and all authors approved the final version. GR, GDW, GG, MYA, KJS, DLG, and KG cared for the patients, devised the treatment plans, and assisted in compiling the clinical histories. JC, MCK, and LDM helped with the processing and interpretation of mass spectrometry data. EEM, NPS, and MK performed the experiments.

## Agreement to Share Publication-Related Data and Data Sharing Statement

For any original data requests beyond what is included in the supplementary material, please email the corresponding author.

## References

1. Pavord S, Scully M, Hunt BJ, et al. Clinical Features of Vaccine-Induced Immune Thrombocytopenia and Thrombosis. N Engl J Med. 2021;385(18):1680–1689.

2. Kanack AJ, Singh B, George G, et al. Persistence of Ad26.COV2.S-associated vaccine-induced immune thrombotic thrombocytopenia (VITT) and specific detection of VITT antibodies. Am J Hematol. 2022;97(5):519-526.

3. Kanack AJ, Schaefer JK, Sridharan M, et al. Monoclonal gammopathy of thrombotic/thrombocytopenic significance. Blood. 2023;141(14):1772–1776.

4. Kanack AJ, Leung N, Padmanabhan A. Diagnostic Complexity in Monoclonal Gammopathy of Thrombotic Significance. N Engl J Med. 2024;391(20):1961–1963.

5. Warkentin TE, Kelton JG. Temporal aspects of heparin-induced thrombocytopenia. N Engl J Med. 2001;344(17):1286–1292.

6. Kohlhagen M, Dasari S, Willrich M, et al. Automation and validation of a MALDI-TOF MS (Mass-Fix) replacement of immunofixation electrophoresis in the clinical lab. Clin Chem Lab Med. 2020;59(1):155–163.

7. Kanack AJ, Bayas A, George G, et al. Monoclonal and oligoclonal anti-platelet factor 4 antibodies mediate VITT. Blood. 2022;140(1):73–77.

8. Wang JJ, Armour B, Chataway T, et al. Vaccine-induced immune thrombotic thrombocytopenia is mediated by a stereotyped clonotypic antibody. Blood. 2022;140(15):1738–1742.

9. Roberge G, Carrier M. Long VITT: A case report. Thromb Res. 2023;223:78–79.

10. Roberge G, Cote B, Calabrino A, Gilbert N, Gagnon N. Acute lower limb ischemia caused by vaccine-induced immune thrombotic thrombocytopenia: focus on perioperative considerations for 2 cases. Thromb J. 2022;20(1):38.

11. Schonborn L, Thiele T, Kaderali L, Greinacher A. Decline in Pathogenic Antibodies over Time in VITT. N Engl J Med. 2021;385(19):1815–1816.

12. Schonborn L, Thiele T, Kaderali L, et al. Most anti-PF4 antibodies in vaccine-induced immune thrombotic thrombocytopenia are transient. Blood. 2022;139(12):1903–1907.

13. Salmasi G, Murray DL, Padmanabhan A. Myeloma Therapy for Monoclonal Gammopathy of Thrombotic Significance. N Engl J Med. 2024;391(6):570–571.

14. Wang JJ, Warkentin TE, Schonborn L, et al. VITT-like Monoclonal Gammopathy of Thrombotic Significance. N Engl J Med. 2025.

