## Supplementary Appendix for "Platelet Factor 4 Antibody Persistence and Long-term Pathogenicity in Vaccine-induced Immune Thrombotic Thrombocytopenia"

**Supplementary Table S1**

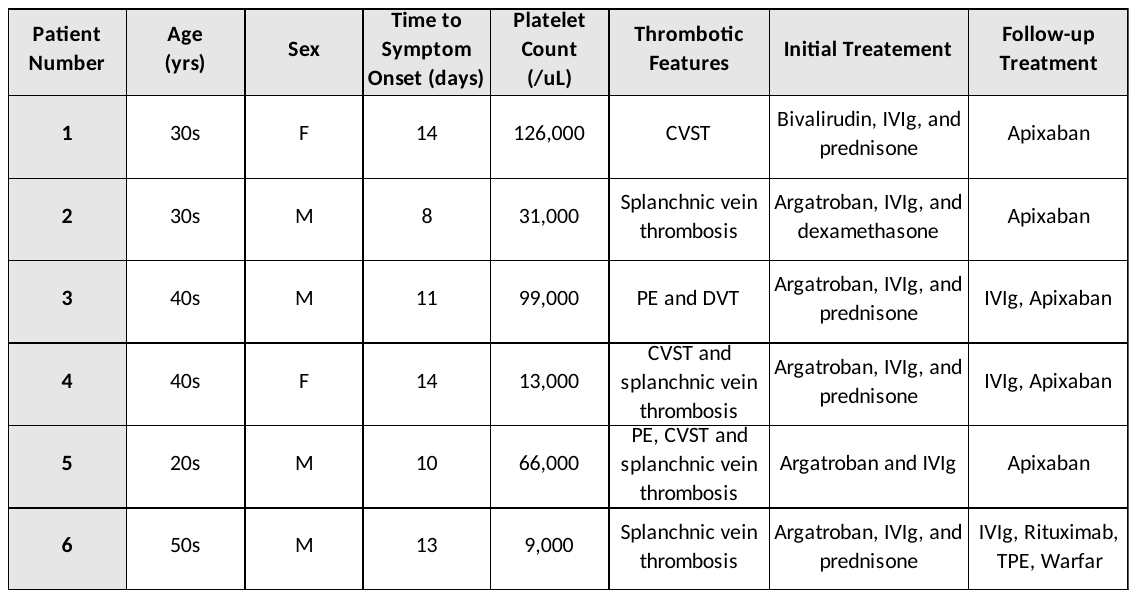

**Table S1. VITT patient demographics and clinical features.** Abbreviations: PE- Pulmonary embolism; CVST- Cerebral venous sinus thrombosis; DVT- Deep venous thrombosis; IVIg- Intravenous Immunoglobulin G.

**Supplementary Table S2**

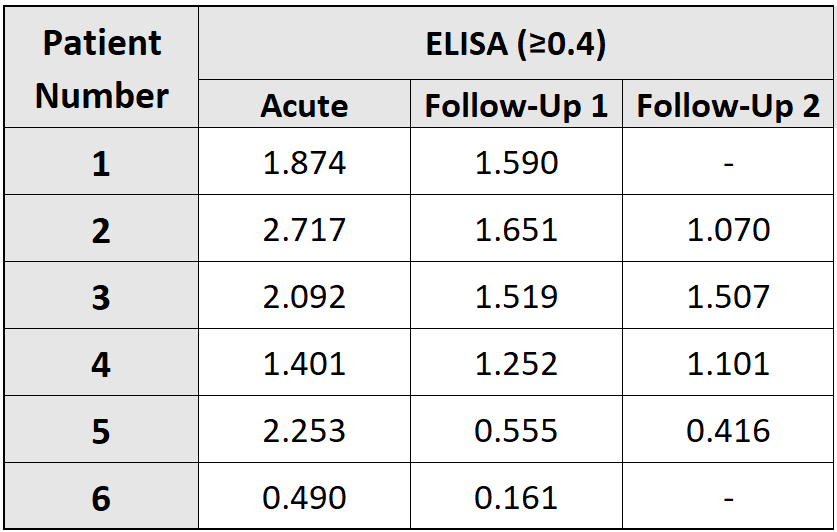

**Table S2. PF4/Polyanion ELISA testing results of acute and follow-up patient samples.** PF4/Polyanion ELISA testing was performed using PF4 IgG ELISA (Immucor). Positive samples were those with an optical density (OD) ≥ 0.400.

**Supplementary Figure S1**

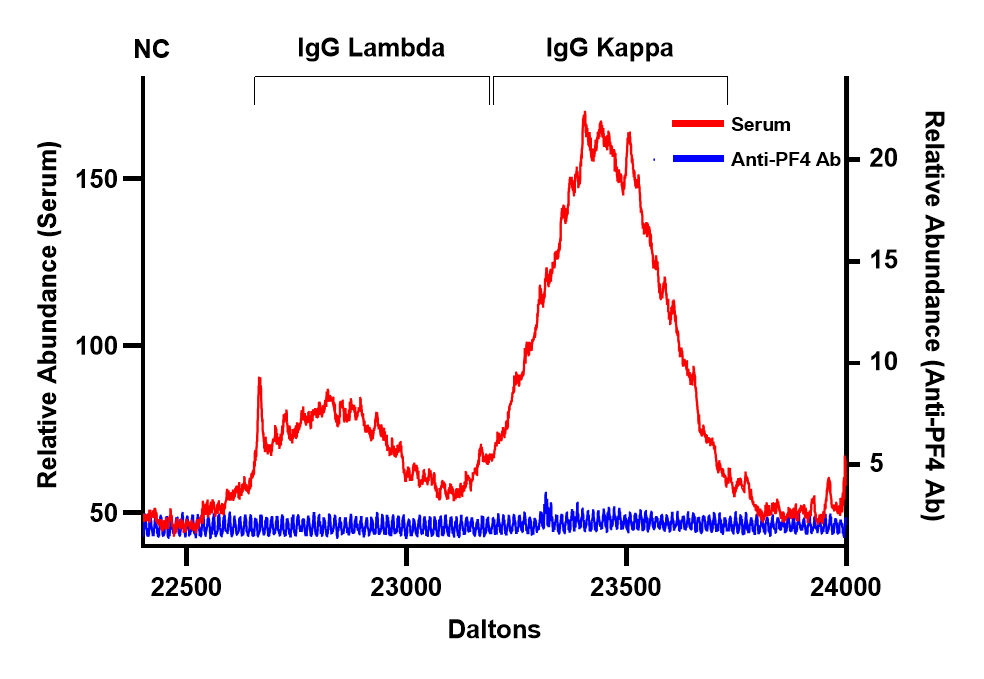

**Figure S1. PF4:Heparin-Sepharose antibody isolations are specific for anti-PF4 antibodies.** LC-ESI-QTOF MS calculated mass spectra (Daltons), based on [M + 12H]^12+^ light chain charge state associated with IgG heavy chains of native serum (red), or PF4:heparin-sepharose bead eluate (blue) from normal donor serum is shown. A mass range of 22400 – 24000 Daltons is displayed on the X axis.

**Supplementary Figure S2**

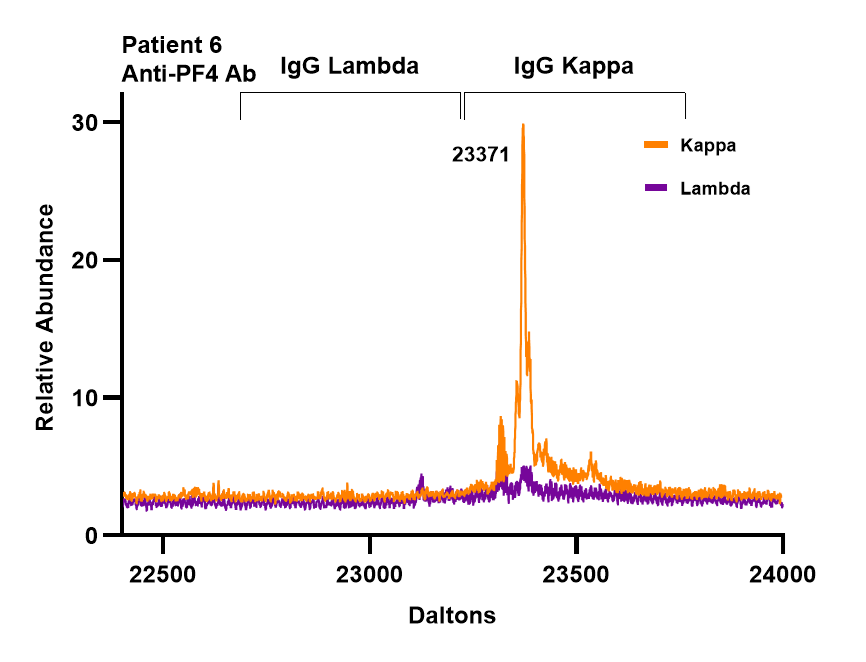

**Figure S2. Anti-kappa and anti-lambda light chain immunoaffinity studies confirm the production of an anti-PF4 kappa light chain by Patient 6.** LC-ESI-QTOF MS calculated mass spectra (Daltons), based on [M + 12H]^12+^ light chain charge state isolated from Patient 6 using PF4-treated heparin sepharose beads is presented. Light chain distributions associated with kappa containing immunoglobulins (Igs) (*Kappa*) and lambda-containing Igs (*Lambda*) are shown in orange and purple, respectively. Values on the X axis represent the mass of the most prominent immunoglobin light chains. A mass range of 22400 – 24000 Daltons is displayed.

**Supplementary Figure S3**

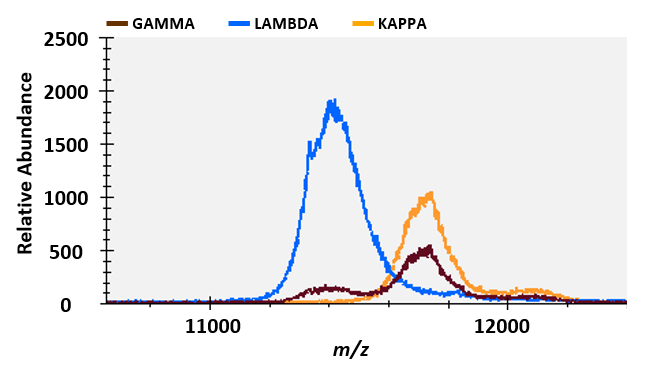

**Supplementary Figure S3. The patient’s monoclonal gammopathy was not detectable in evaluation of total IgG repertoire in patient serum by matrix-assisted laser desorption ionization time-of-flight mass spectrometry analysis (“Mass-Fix”).** Shown are Mass-Fix +2 spectra of separate immune enrichments for IgG heavy chain (GAMMA) in brown, *lambda* containing Igs (LAMBDA) in blue, and *kappa* containing Igs (KAPPA) in orange. The x-axis depicts mass-charge (*m/z*), and the y-axis presents the relative abundance of identified antibodies.

**Supplementary Figure S4**

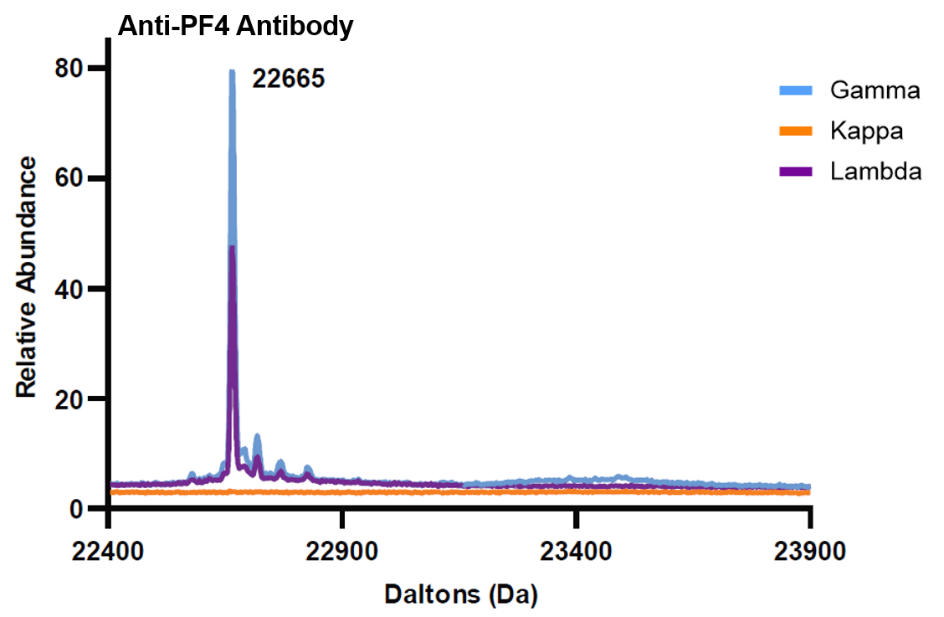

**Supplementary Figure S4. Patient’s anti-PF4/polyanion antibody isolation displays near-identical mass spectrometry profile to antibody eluted from PF4-coupled beads.** LC-ESI-QTOF MS deconvoluted charge state spectra (Daltons) of antibodies isolated using PF4-treated heparin sepharose beads. Light chain distributions associated with an IgG heavy chain (*Gamma*) in blue, kappa containing Igs (*Kappa*) in orange, and lambda containing Igs (*Lambda*) in purple are shown. Values on the x-axis represent the mass (Da) of the most prominent immunoglobin light chain.

**Methods**

*Patient Samples*

Patient serum samples were obtained from six individuals with Ad26.COV2.S-associated VITT and one ChAdOx1 nCov-19-associated VITT. Research studies were approved by the Institutional Review Board of Mayo Clinic.

*Functional platelet studies*

The PF4-dependent P-selectin expression assay (PEA) was performed as previously described.^1, 2^ Prostaglandin E1 was added to citrated whole blood obtained from healthy volunteers to a 50 ng/mL concentration. Whole blood was centrifuged at 200 x g for fifteen minutes to obtain platelet-rich plasma (PRP). Platelet isolation was performed by centrifugation at 1,000 x g for ten minutes and resuspension of the platelet pellet in phosphate-buffered isotonic saline (pH 7.4) with 1% bovine serum albumin (PBS-BSA). Platelets (1 x 10^6^) were then treated for twenty minutes at ambient temperature with PF4 (37.5 μg/mL) or PBS. Ten microliters of the patient sample and forty microliters of platelets were incubated for one hour at ambient temperature before the addition of fluorescently labeled anti-P-selectin (monoclonal antibody HB-299, ATCC) and anti-GPIIIa (monoclonal antibody HB-242, ATCC) antibodies for twenty minutes. A final volume of 200 µL was then obtained by the addition of PBS-BSA, and platelet events were gated based on GPIIIa positivity before P-selectin expression (median fluorescence intensity, MFI) was recorded.

*PF4-Polyanion HIT ELISA*

PF4 IgG (Immucor) ELISA was used according to the manufacturer’s instructions. In brief, patient serum was incubated with PF4-PVS coated platelets, washed, and incubated with an alkaline phosphatase-labeled anti-human IgG antibody. After antibody incubation, p-nitrophenyl phosphate (pNPP) substrate was added, and colorimetric detection was performed at optical densities of 405 and 492nm after thirty minutes.

*VITT ELISA*

Detection of VITT/VITT-like anti-PF4 antibodies was performed using uncomplexed, chemically cross-linked recombinant PF4. PF4 cross-linkage was completed using equal ratio (v:v) of EDC (1 mg/mL) and recombinant PF4 in 15 mM MES buffer pH 6.0 for six hours at ambient temperature. Reactions were halted using 1M Tris pH 7.4 buffer at a 1:4 (v:v) ratio of Tris buffer to cross-linked PF4 polypeptide reaction. Uncomplexed cross-linked PF4 (0.5 µg/well) was then plated on ELISA plates (Thermo Scientific) overnight at 4°C. ELISA plates were washed three times with PBS pH 7.4 +0.1% TWEEN®-20 and blocked with SUPERBLOCK T20 (Thermo Scientific). Following dilution (1:50) patient samples were added then incubated for an hour, followed by four washes with PBS/0.1% TWEEN®-20. Alkaline phosphatase-conjugated goat anti-human IgG Fc antibody (Jackson Immunoresearch) was added at a dilution of 1:5000. Four additional washes were performed using PBS/0.1% TWEEN®-20 and colorimetric detection was performed using p-nitrophenyl phosphate (pNPP) substrate. The optical density (OD; 405 nm minus 492 nm) at 30 minutes was recorded. The positive cut-off of the VITT ELISA was set at three standard deviations above the mean optical density of 50 healthy donor samples (0.341).

*Antibody Isolation*

Anti-PF4 antibodies were isolated using PF4-coupled MyOne Carboxylic Acid Dynabeads (500μL, ThermoFisher) as previously described^4^. Briefly, beads were washed with 15mM MES pH 6.0 and then activated using 50μL of 1-Ethyl-3-(3dimethylaminopropyl) (EDC, 10mg/mL) for 30 minutes. The beads were again washed and then incubated with 250μg of PF4 in MES overnight. Following washing with PBS/0.1% Tween-20, beads were blocked with PBS/0.1% BSA for 10 minutes. Then 250μL of patient sample (serum) was incubated with the beads for 2 hours. Beads were thoroughly washed with PBS, and elution of the antibodies using 100 mM glycine pH 11.0 for 5 minutes was completed. Eluted antibodies were neutralized with 1M Tris and dialyzed against PBS before being evaluated by HIT ELISA, PEA, and mass spectrometric studies.

Anti-PF4 antibodies were also isolated as described recently^3^. Heparin sepharose beads (200 μL, Cytiva Lifesciences) were washed with PBS, pH 7.4, and incubated with 200 μg of recombinant PF4 for one hour. Five hundred microliters of patient samples were added to beads for one hour. Beads were thoroughly washed with PBS, and eluates were obtained from the PF4/heparin sepharose beads using 2M NaCl. Eluates were dialyzed against PBS before being evaluated by HIT ELISA, PEA, and mass spectrometric studies.

*Serum protein electrophoresis*

Serum protein electrophoresis was performed according to protocols in the Clinical Immunology Laboratory, Department of Laboratory Medicine and Pathology, Mayo Clinic, on the SPIFE 3000 electrophoresis analyzer (Helena Laboratories). The total protein concentration was determined by colorimetric assay using reagents from Siemens and a Siemens Advia Chemistry XPT system (Siemens Medical Solutions USA, Inc).

*Liquid Chromatography Electrospray Ionization time-of-flight mass spectrometry*

The basic method used for antibody analysis has been previously described.^4, 5^ Immunoglobulins (Igs) from patient sera or bead eluates were isolated using camelid-derived nanobodies selective for the constant domains of human Ig gamma heavy chain, kappa light chain, or lambda light chains (CaptureSelect affinity resins, Thermo Fisher Scientific). One hundred microliters or fifty microliters of camelid nanobody beads were incubated with ten microliters of serum or one hundred microliters of anti-PF4 antibody eluate, respectively, diluted with two hundred microliters of PBS and incubated for thirty minutes at ambient temperature. Subsequently, sera or eluate supernatants were removed from the beads, and the beads were washed three times with five hundred microliters of water. Ig from light and heavy chains of sera and anti-PF4 antibody eluate were eluted using sixty microliters or twenty microliters of 5% acetic acid, respectively. Following a five-minute incubation, eluted Igs were reduced using 100 mM dithiothreitol (DTT) in 1M ammonium bicarbonate (2:1; v:v) to disassociate immunoglobulins and to separate light chain and heavy Ig chains. An Agilent 1290 Infinity II liquid chromatography (LC) system was used to separate Ig chains before ionization and to remove any co-eluted PF4 before analysis using a SCIEX Zeno time-of-flight (TOF) 7600 mass spectrometer (MS). Ten microliters of each camelid nanobody bead eluate was injected per analysis onto a Poroshell 300SB-C3 column (2.1 mm X 75 mm) with a 5 μm particle size placed in a 60 ºC column heater. The mobile phases included an aqueous phase A (100% water + 1% formic acid) and an organic phase B (90% acetonitrile + 10% isopropanol + 0.1% formic acid), and the flow rate was 300 μL/min. The light chains elution range was identified during a 4.5-minute gradient from 27%B to 32%B. The diverter valve was used to direct 10.35 minutes of the gradient into the MS; otherwise, the LC was diverted to waste. The MS, using positive electrospray ionization, was run using intact protein workflow; CUR 30, CAD 7 GAS1 35, GAS2 30, and temperature 500 °C. TOF MS data from collected from 600 to 2500 m/z; DP 175 and CE 10. Data analysis was performed using Sciex OS v2.2 and PeakView ver. 2.2. Mass spectra (Daltons) was calculated based on [M + 12H]^12+^ light chain charge state as described elsewhere.^4, 5^ The retention time of the monoclonal light chain in each patient sample was tracked using PeakView. The mass spectra of the multiply charged light chain ions were deconvoluted to obtain an accurate molecular mass using the Bio Tool Kit ver. 2.2 plug-in software.

*Matrix-assisted laser desorption ionization time-of-flight Mass Spectrometry*

Mass-Fix was performed on patient samples as previously described^6^. CaptureSelect resins (Thermo Fisher) were washed with 30 volumes of PBS + 0.1% Tween-20. Using a final working volume of resins of 10% w/v, fifty microliters of each CaptureSelect Resin solution was pipetted into the wells of a 384-well plate, and ten microliters of patient serum were added to each well. After incubating for fifteen minutes, the serum supernatant was removed from the resin. The resin was washed using 50 microliters of PBS three times and 50 microliters of water three times. Captured Igs were then eluted and reduced using 30 mcl of 20 mM TCEP + 0.10% TFA. Finally, sample eluates were diluted 1:2 in a separate dilution plate, using TCEP + 0.10% TFA. A ttpLabtech Mosquito nanoliter pipettor (Hertfordshire, United Kingdom) was utilized for spotting. Sample eluate (0.5 microliters of purified patient Igs in 0.1% TFA containing ten mM TCEP) was combined with alpha-cyano-4-hydroxycinnamic acid (CHCA) matrix (0.7 microliters, 10 mg/mL in 50% ACN + 0.1% TFA) and spotted onto a 96-well microScout polished steel Bruker target (Bruker Daltonics) using a single application spotting method. Analysis was performed in positive ion mode with a summation of 500 laser shots using a MALDI-TOF mass spectrometer (Bruker Microflex LT, Germany).

**References**

1.	Padmanabhan A, Jones CG, Curtis BR, Bougie DW, Sullivan MJ, Peswani N, McFarland JG, Eastwood D, Wang D, Aster RH. A Novel PF4-Dependent Platelet Activation Assay Identifies Patients Likely to Have Heparin-Induced Thrombocytopenia/Thrombosis. Chest. 2016;150(3):506-15. Epub 20160219. doi: 10.1016/j.chest.2016.02.641. PubMed PMID: 26905366; PMCID: PMC5028397.

2.	Samuelson Bannow B, Warad DM, Jones CG, Pechauer SM, Curtis BR, Bougie DW, Sharma R, Grill DE, Redman MW, Khalighi PR, Leger RR, Pruthi RK, Chen D, Sabath DE, Aster RH, Garcia DA, Padmanabhan A. A prospective, blinded study of a PF4-dependent assay for HIT diagnosis. Blood. 2021;137(8):1082-9. Epub 2020/09/09. doi: 10.1182/blood.2020008195. PubMed PMID: 32898858; PMCID: PMC7907721.

3.	Kanack AJ, Bayas A, George G, Abou-Ismail MY, Singh B, Kohlhagen MC, Splinter NP, Christ M, Naumann M, Moser KA, Smock KJ, Grazioli A, Wen R, Wang D, Murray DL, Padmanabhan A. Monoclonal and oligoclonal anti-platelet factor 4 antibodies mediate VITT. Blood. 2022;140(1):73-7. Epub 2022/05/14. doi: 10.1182/blood.2021014588. PubMed PMID: 35560046; PMCID: PMC9262283.

4.	Barnidge DR, Dasari S, Ramirez-Alvarado M, Fontan A, Willrich MA, Tschumper RC, Jelinek DF, Snyder MR, Dispenzieri A, Katzmann JA, Murray DL. Phenotyping polyclonal kappa and lambda light chain molecular mass distributions in patient serum using mass spectrometry. J Proteome Res. 2014;13(11):5198-205. Epub 20140826. doi: 10.1021/pr5005967. PubMed PMID: 25134970.

5.	Barnidge DR, Dasari S, Botz CM, Murray DH, Snyder MR, Katzmann JA, Dispenzieri A, Murray DL. Using mass spectrometry to monitor monoclonal immunoglobulins in patients with a monoclonal gammopathy. J Proteome Res. 2014;13(3):1419-27. Epub 20140211. doi: 10.1021/pr400985k. PubMed PMID: 24467232.

6.	Kohlhagen M, Dasari S, Willrich M, Hetrick M, Netzel B, Dispenzieri A, Murray DL. Automation and validation of a MALDI-TOF MS (Mass-Fix) replacement of immunofixation electrophoresis in the clinical lab. Clin Chem Lab Med. 2020;59(1):155-63. Epub 20200803. doi: 10.1515/cclm-2020-0581. PubMed PMID: 32745067.
